# Systematic literature review examining the mere-measurement effect of patient reported measures: the interaction of emotional valence and frequency of exposure as an independent factor in patient change

**DOI:** 10.1101/2022.03.29.22273094

**Authors:** Preston Long, Valentin Ritschl, Lisa Otto, Alize Rogge, Linetta Koppert, Anouk Huberts, Tanja Stamm

## Abstract

The use of patient-reported outcome (PROs) have become increasingly commonplace across many healthcare settings over the past two decades. The value of PROs is now acknowledged by healthcare providers and patients alike. However, to date, little is known about the best practices for formulating PRO measures (PROMS), but even more specifically, the effect had on the responding patients as a result of item word choice, emotional valence, or frequency of use. That is, 1) does the positive or negative wording of items affect the patient’s perspective on the latent variable, 2) is there a degree of subliminal influence or measurement effects on their behaviour resulting from exposure to PROs, and finally, 3) is such an effect amplified with repeated exposure?

## Background

There is currently sufficient evidence to suggest that attributes such as the wording of questions, their presentation order, the context where they are asked, and the item’s social sensitivity (e.g. I do not abuse my prescriptions), have an effect on participant responses (Schwarz, 1999; Näher & Krumpal, 2012). This family of variables are most oft referred to as method effects. Method effects largely regard the phenomenon in which the presentation of an item affects the participant’s response independent of the content in question. The end goal within this area of study is to correct for any response bias resulting from the method used. This research primarily concerns psychometrics. It is important to note that while method effects relate to the research questions proposed, it is not the key feature of interest. The ensuing inquiry will not simply focus directly on changes to item responses, but instead, on changes to patient perceptions and behaviours as a result of their exposure to the measures themselves.

Research supports the claim that asking certain questions can influence behaviours on the same topic (Sandberg & Conner, 2009; Godin et al., 2010). This phenomenon is called the mere-measurement effect. For example, a randomized-control trial was conducted to assess the impact of asking questions pertaining to blood donation habits of the participants. The researchers found that the subjects whom were asked about their habits were significantly more likely to donate blood than their control counter parts at the following blood-drive (Godin, Sheeran, Conner, & Germain, 2008). Other researchers found that when clinicians did not ask patients with Attention Deficit Hyperactivity Disorder (ADHD) to report whether they were currently medicated on a disease-related severity questionnaire, patients were more likely to report un-medicated symptomologies (Lineweaver et al., 2021). In other words, patients on ADHD medications responded more symptomatically similar to their un-medicated selves when their medication status was not included on the questionnaire. This suggests that the medication status item itself, may have mitigated ADHD symptoms by bringing the patient’s treatment into a higher state of awareness/directing attention.

Thus, it may be that asking patients about their intent to have an operation increases the likelihood of them opting for it, or, that asking them about their exercise increases their physical activity level. If so, this may have significant implications for the effect of PROs, as well as for utilizing measurements as interventions. However, before value can be attached to such an effect, first the presence and strength must be evaluated in the patient-specific context.

To date, almost no method effects and even less mere-measurement effect factors have been investigated in the context of PROMs and particularly not in regard to how they may influence patient perceptions and behaviours. Therefore, exploration of this family of effects must be reduced to a few key anticipated features, namely, emotional valence (positive/negative) and frequency. For instance, would wording a cleft-lip question negatively, such as “I do not like my face”, actually decrease the patient’s preference for their own face, and, does repeated exposure to this question increase the magnitude of the effect.

Due to the degree of overlap between method and mere-measure effects, and the anticipated paucity of scholarly works on the matter, method effects must also be included albeit with their appropriateness closely scrutinized. This systematic literature review will be the first to the authors’s knowledge to examine if measuring patient outcomes directly effects their corresponding behaviour, and if this effect can be attributed in part to item wording and/or frequency of exposure.

## Aim

The aim of this SLR is to discover, collect, analyse, and report what is currently known about the effects of measuring patient reported outcomes, as it relates to patient perceptions and behaviours on the item topics of inquiry.

Research Questions:

1. Does the assigned positivity or negativity of an item effect patient perception or behaviour on the topic inquired?
2. Does frequency of exposure to an item have an effect on patient perception or behaviour of the topic inquired?

## Method

The following tasks will be performed within this project (van der Heijde et al., 2014)

1. Establishing a steering committee including experts in the field of outcomes research and psychology
2. Having a first steering committee meeting to define scope of study, keywords, MeSH-Terms and a search strategy for the SLR
3. Conducting the SLR supervised by the methodologist
4. Reviewing and discussing the findings of the SLR in a second steering committee meeting
5. Reporting the results according to the Preferred Reporting Items for Systematic Reviews and Meta-Analyses (PRISMA) statement.

### Members of the international steering committee

The steering committee will include …. TS, VR, PL, LO, AR

### First (virtual) steering committee meeting to define research questions, keywords, MeSH-Terms and search strategy for the SLR

The core team (coordinating investigators and methodologist) will draft the search question(s), identify relevant keywords, MeSH-Terms and develop the search strategy for the SLR on different databases and refine these based on the feedback from the steering committee members. The research question(s) will then be converted into the PICO (Patient, Intervention, Comparison, Outcome) scheme by the core team. Within the steering committee, it will also be discussed, whether to limit the SLR to certain assessments and treatment options.

PICO Questions:

In an adult population of patients*, does exposure to a concept or topic on an instrument effect the patient’s perspective or behaviour on that respective topic?

In an adult population of patients*, does the emotional valence of an item’s wording, compared to neutral wording, effect the patient’s perspective or behaviour on the topic of the item?

In an adult population of patients*, does repeated exposure to the same topic have a mounting effect on the patient’s behaviour or perceptions over time? And, is this effect consistent across emotional valences?

*defined as people completing the study in a healthcare setting and/or responding to items about their illness status (i.e. treatment, recovery, etc.), or healthcare decisions (including hypotheticals and vignettes).

#### Operational definition of PROs

For the purposes of this review, patient-reported outcomes are defined as outcomes derived directly from subjective participant input which is collected in the context (physically or hypothetically) of healthcare, and/or, the implemented questionnaire’s focus is directly related to a responder’s clinical diagnoses.

### Performing the SLR to collect the evidence

A SLR based on the identified PICO questions will be undertaken by the methodologist (PL) and a research fellow (TS). Relevant records will be identified using the databases: MEDLINE [PubMed], CINAHL [Ebsco], Web of Science, ScienceDirect, and the Cochrane Database of Systematic Reviews. The following study designs will be included: systematic reviews (SR), randomised controlled trials (RCT), controlled trials (CT), quasi-experimental studies, observational studies, cohort studies, case-control and qualitative studies. There will be no exclusions based on publication date. All results obtained by the researchers conducting the SLR will be discussed with the coordinating investigators and the steering committee in a following step. The platform Covidence will be used for the search, title and abstract screening, and full-text review.

### Proposed draft search strategy for the SLR

We will search in MEDLINE [PubMed], CINAHL [Ebsco], Web of Science and ScienceDirect. The search will be based on PsyInfo-Terms and keywords regarding questionnaires, surveys, method effects, word choice, emotional valence (positivity or negativity), mere-measurement effects, mere-exposure effects, item effects, and patient outcomes. Method effects will be included in the search because of the dearth of research in this area and the overlap between it and mere-measurement. Due to the strong focus on patients in this SLR, it is possible to search with the following PsyInfo-Terms: “Patient, behaviour”[Mesh] AND (“Mere-measurement, Patients “[Mesh] OR “Patient method effects”[Mesh]). An extensive, more detailed list of keywords will be used for data extraction and further analyses. The search strategy exemplified on PubMed is displayed in table 1.

**Table 1.**
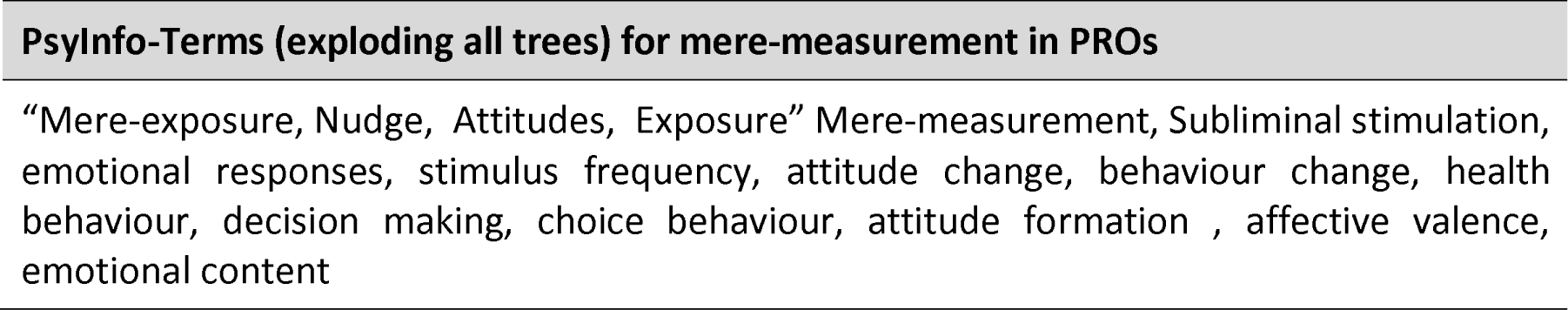
Search strategy exemplified on PsyInfo

Subsequently, steering committee members will be consulted by email to identify additional grey literature or research that might not be found through the process mentioned above. Furthermore, reference lists of identified papers and existing reviews will be searched to select relevant articles subject to the same screening and selection process as original/initial papers.

### In- and exclusion criteria

After gathering the evidence, the eligibility criteria (listed below) will be applied to the results. All identified references will be screened independently by two reviewers (PL and TS from the Medical University of Vienna and LO and AR from Charite’). The selection process will consist of two phases. A first screening will be performed on title and abstract whether they met inclusion/exclusion criteria or not. In a second step, the eligibility of the full text articles will be explored.

When in doubt whether a paper meets the inclusion criteria, the coordinating investigator VR will be asked to assess the paper and the decision will be made by consensus.

#### Inclusion criteria

- This SLR will focus on identifying perceptual or behavioural changes in patients as a result of exposure to questionnaire items
- Studies will be included in this SLR if participants in the studies are overtly considered patients, i.e. have a reported diagnosis
- Studies will be eligible if they sufficiently focus on the outcomes of patients
- All settings will be taken into account (home care, community services, primary health care, hospital settings, laboratory etc.)
- All participants will be included regardless of demographics (i.e. age, race)
- The following types of studies will be included: systematic reviews (SR), randomised controlled trials (RCT), controlled trials (CT), quasi-experimental studies, observational studies, cohort studies, case-control studies and qualitative studies
- There will be no exclusions based on publication date

#### Exclusion criteria

- Mere-measurement studies conducted on non-patient participants
- Papers describing the development of new psychometric methods, assessments, or screening tools
- Non-human participants
- Participants less than 18 years of age
- Studies will be excluded if they are not written in English
- Studies that focus on clinical or physiological outcomes
- Studies must include a follow-up after the initial measurements

### Data collection and analysis, assessment of included studies, risk of bias assessment and synthesis of evidence

Data extraction will be based on the predefined inclusion and exclusion criteria and performed by PL, AH, LO, and AR using pilot-tested data extraction forms. The quality of all retrieved papers will be assessed by two reviewers (TS & LK) using validated assessments tools. VR will act as the methods supervisor and ensure extraction quality. Systematic reviews will be critically appraised using A MeaSurement Tool to Assess Systematic Reviews (AMSTAR 2).

For each question, a narrative summary will be undertaken to report on the study characteristics. Tables will accompany the narrative summary, to ensure the consistency of the presented information across all studies and facilitate study comparisons by the reader. A narrative summary of the results of the critical appraisals will be presented separately for each research question, including an overall impression of the quality of included studies. If there is ample primary data a meta-analysis will also be conducted.

### Reviewing and discussing the findings of the SLR in a second (virtual) steering committee meeting

In this meeting, a framework for developing and presenting summaries of evidence and providing a systematic approach for making both applied recommendations and theory-based inferences will be created.

### Reporting the results

The results of the SLR will be published according to the Preferred Reporting Items for Systematic Reviews and Meta-Analyses (PRISMA) statement in a peer-reviewed journal.

## Data Availability

All data produced are available online

